# Understanding Low Vaccine Uptake in African, Caribbean, and Black Populations Relative to Public Health in High-Income Countries: A Scoping Review Protocol

**DOI:** 10.1101/2024.01.14.24301294

**Authors:** Josephine Etowa, Sheryl Beauchamp, Victoria Cole

## Abstract

**Background:** Vaccination has significantly contributed to reducing once common and even deadly infectious diseases, yet vaccine hesitancy threatens the emergence of vaccine-preventable diseases. The COVID-19 pandemic has caused the need for the largest mass vaccination campaign ever undertaken to date; however, African, Caribbean, and Black (ACB) populations have shown both a disproportionately high degree of negative impacts from the pandemic and the lowest willingness to become vaccinated. Low vaccination rates in this vulnerable population are a pinnacle concern in public health, as low vaccination rates in ACB communities may both be the result of health inequities, as well as further exacerbate them.

**Purpose:** To explore low vaccine uptake in African, Caribbean, and Black (ACB) populations relative to public health in high-income countries. *Objectives* 1) To identify concepts and boundaries of existing evidence sources on low vaccine uptake in ACB populations; 2) To map out the evidence on the concepts and boundaries and to identify gaps in the research; and 3) To determine existing interventions to improve low vaccine uptake in the study population.

**Methodology:** This scoping review follows the Joanna Briggs Institute (JBI) framework for scoping reviews, supplemented by the Preferred Reporting Items for Systematic Reviews extension (PRISMA-ScR). Any deviations from the JBI recommendations are stated. ***Theoretical underpinnings*** of the intersectionality approach will be used to help interpret the complexities of health inequities in the ACB population, including those related to the social determinants of health (SDOH). ***Search strategies*** were developed by an information specialist (VC) and peer-reviewed using the PRESS guideline. The search was conducted in: MEDLINE(R) ALL (OvidSP), Embase (OvidSP), CINAHL (EBSCOHost), APA PsycInfo (OvidSP), Cochrane Central Register of Controlled Trials (OvidSP), Cochrane Database of Systematic Reviews (OvidSP), Allied & Complimentary Medicine Database (Ovid SP), and Web of Science. ***Eligibility criteria*** are based on the Population, Concept, Context (PCC) framework. The inclusion criteria for this study included evidence -sources with a primary focus on African, Caribbean, and Black populations, and other related terms; high-income countries as defined by the World Bank where ACB populations are considered a minority; all service providers; English and French languages; all types of evidence sources; related to low vaccine uptake and alternative terms; all vaccines; and publications from 2020- current (July 19^th^, 2022). ***The screening, selection, and extraction*** of the evidence sources were completed by the AVA research team. ***Analysis*** was done through the process of Thematic Mapping, and ***summarization and presentation*** of the findings were done through a narrative description organized using the socioeconomic model (SEM) as a framework.

**Ethics and dissemination:** This study used published evidence sources with no human or animal participants; thus, ethical approval and consent to participate are not applicable.

**Dissemination:** This will occur through peer-reviewed open-access journals and conferences that target stakeholders in public health, vaccination campaigns and overcoming inequities in healthcare.

## Introduction

African, Caribbean, and Black (ACB) populations are not only vulnerable due to health inequities, as evidenced by higher rates of SARS- CoV-2 infections, hospitalizations, and associated mortalities, but are also the least willing to receive the vaccine.[1–3] The COVID-19 pandemic has been one of the greatest public health threats of modern times, bringing societal, community, and individual challenges to the forefront; these impacts have been intensified in racialized communities, as pre-existing inequities and vulnerabilities are exacerbated.[4,5] These inequities are strongly influenced by socioeconomic factors, referred to as the social determinants of health (SDOH); for example, death rates in ACB populations were higher in areas with a greater incidence of adverse SDOH.[5,6] Although race-based data collection remains inconsistent in Canada, the cities of Ottawa and Toronto reported 1.5-5 times the increase in COVID-19 infection rates among racialized communities; these findings are consistent with other high-income countries, including the United States (US) and the United Kingdom (UK).[5,7,8] These highlighted COVID-19 racial inequities regarding disease and vaccinations in ACB populations, are not new; public health disparities also occurred during the 2009 H1N1 pandemic, and high rates of vaccination mistrust have been reported for the Human Papillomavirus (HPV) vaccine, H1N1 vaccine, and influenza vaccine.[4,9–12] The World Health Organization (WHO)[13] defines vaccine hesitancy as the refusal or reluctance to become vaccinated despite available vaccines; the reasons identified include complacency, inconvenience, and lack of confidence. It can result in delayed vaccination or uncertainty in the vaccine even after its administration, which can threaten vaccination programs by leading to decreased coverage and increased risk of vaccine-preventable disease outbreaks.[14,15] Currently, vaccine hesitancy regarding the COVID-19 vaccination threatens the success of the largest mass vaccination campaign ever undertaken to date, both within Canada and globally.[16] However, the importance of addressing vaccine hesitancy reaches beyond its implications for COVID-19, including both current and potential future outbreaks; the coronavirus alone has accounted for three pandemics within the last 20 years, including COVID- 19, severe acute respiratory syndrome (SARS), and Middle East respiratory syndrome (MERS).[17]

Vaccine uptake has been declining over the past several decades; global coverage dropped from 86% in 2019 to 83% in 2020, with the highest rate of children under one, 23 million, not receiving basic vaccines since 2009, and completely unvaccinated children increasing by 3.4 million.[18,19] In 2019, the World Health Organization (WHO) listed both vaccine hesitancy and weak primary healthcare as two of the top ten threats to global health; both threaten the success of vaccination campaigns.[13,20]

In Canada and globally, vaccines significantly prevent and control infectious diseases and are thus a cornerstone of public health.[21] Historically, vaccines have reduced disease-specific mortality rates, including smallpox, rabies, polio, the plague, typhoid and many more, and have significantly decreased infant mortality rates globally[18]; over 3 million child deaths are estimated to be prevented each year globally, through vaccinations.[22] Despite vaccinations being considered to be one of the public health’s greatest success stories, vaccine hesitancy is influenced by the confidence in the competencies of health professionals and health services.[23,24] Vigilance is required to maintain and increase vaccine uptake, especially in vulnerable populations, as people’s behaviors and willingness to follow recommended measures are the most powerful tools against viral spread.[25,26]

Public health interventions must go beyond the COVID-19 vaccine and seek to understand the historical basis for vaccine hesitancy, while adapting to the current dynamics and preparing for future outbreaks. If public health fails to implement appropriate interventions, health inequities threaten to become even more vast, as those who are socioeconomically disadvantaged often have health conditions that are exacerbated by inadequate healthcare.[10,27,28] Further research is needed to determine why ACB populations have the lowest level of vaccine acceptance in Canada than other high-income countries.[3,29]. These disparities need to be promptly addressed; however, a greater understanding about the implications of challenges faced by vulnerable populations on vaccine uptake and public health is required.[10,28]

Prior to embarking on creating service provider (i.e., healthcare providers, policymakers, and community organization providers) interventions applicable to vaccine hesitancy in vulnerable ACB populations, associated concepts and their boundaries must be clarified. Due to the explorative nature and broad overview desired, a scoping review (ScR) approach has been chosen. There has been a steady increase in the use of ScRs, as they are valuable for health researchers to establish the breadth of data available.[30,31] Due to this ScR not undergoing assessment of bias, including critically appraising the evidence sources, the implication for service providers would be better served through a systematic review however, if the evidence sources reveal any potential implication to service provider practice, service provider knowledge and research, these will be stated.[32,33] The essential characteristics of this ScR will include pre-planning through the creation of a protocol, transparency of the processes involved, and clarity of concepts.[31]

A preliminary search in JBI Evidence Synthesis, Cochrane Database of Systematic Reviews, Cumulative Index to Nursing and Allied Health Literature (CINAHL) and PubMed, for existing scoping reviews, systematic reviews and protocols was performed on January 31^st^, 2022, relating to the determinants of vaccine hesitancy and ACB populations. This search was done using Several terms (refer to S1 File), and the keywords such as “vaccine hesitancy” and “Black” yielded the most relevant results, namely, the following four reviews: One rapid systematic review related to COVID-19 vaccine hesitancy and minority ethnic groups in the UK[34]; one scoping review related to COVID-19 vaccine hesitancy globally[35]; and two systematic reviews, one related to vaccine hesitancy in the US[36] and the second related to vaccine acceptance in different populations in the US.^38^ In addition, on June 22^nd^, 2022, VC searched the Open Science Framework (OFS) site and found the ScR protocol on racial and ethnic minorities and Indigenous population groups living in high-income countries.[38] To avoid duplication of findings, this ScR included all vaccines, ACB populations specifically, all high-income countries where the ACB population is considered a minority population, and evidence sources from 2020- 2022.

To develop a clear study structure, and to help guide the selection of evidence sources that align with the research question, the Joanna Briggs Institute (JBI) recommended PCC (population, concepts, context) framework was used; PCC was also used to compose the title as per the JBI framework.[32] The research question is: What are the determinants and interventions of low vaccine uptake in African, Caribbean, and Black (ACB) populations relative to healthcare in high-income countries? Whereby the population (P) is ACB populations and service providers, the concept (C) is low vaccine uptake, and the context (C) is healthcare in high income.

The aim of this ScR is to explore low vaccine uptake in ACB populations relative to public health in high-income countries. The objectives are: 1) To identify concepts and boundaries of existing evidence sources on low vaccine uptake in ACB populations; 2) To map the evidence on the concepts and boundaries and to identify gaps in the research; and 3) To determine existing interventions to improve low vaccine uptake in the study population. These objectives were achieved through systematically reviewing the breadth and types of source evidence available.

This scoping review is part of a larger study examining vaccine uptake in the ACB community which has been funded by the Public Health Agency of Canada. Funding number: # 2122-HQ- 000318. The study received full ethical approval from the University of Ottawa Research Ethics Board (REB). University of Ottawa ethics approval certificate number: H-12-21-7558.

## Methodology

The Joanna Briggs Institute (JBI) scoping review (ScR) methodological framework by Peters et al.[32] will help provide organization to this ScR, which aims to outline different types of evidence on the determinants of vaccine hesitancy in ACB populations, and the gaps for future research. The nine steps of the framework are; 1) defining and aligning the objectives and research question; 2) developing and aligning the inclusion criteria with the objectives and question; 3) describing the planned approach to evidence searching, selection, data extraction and evidence presentation; 4) searching the evidence; 5) selecting the evidence; 6) extracting the evidence; 7) analyzing the evidence; 8) presenting the evidence; and 9) summarizing the evidence in relation to the purpose of the review, making conclusions, and noting any implications of the findings.[32]

### Developing and Aligning the Inclusion Criteria with the Objectives and Questions

The inclusion criteria are: evidence-sources with a primary focus on African, Caribbean, and Black (ACB) populations and related terms; high-income countries as defined by the World Bank[39] where ACB populations are considered a minority; all service providers; English and French languages; all types of evidence sources; related to low vaccine uptake and alternative terms; all vaccines; and publications from 2020-current (July 19^th^, 2022).

The exclusion criteria are: Evidence sources prior to 2020, racialized and minority populations that are not ACB, and any evidence source that does not meet the inclusion criteria.

### Eligibility Criteria with Explanation in PCC Framework

The population (P) includes any person, community, or population that is identified as African, Caribbean, and Black (ACB) or related terms. This population has been chosen because of their high levels of vulnerability and high propensity for vaccine hesitancy. The evidence source must describe the population in a high-income country. This is to aid in exploring the roles of race and ethnicity on the health of ACB populations in Canada, which has often been extrapolated from US and UK-derived statistics; two countries that systematically collect race-based health data. The case of collecting race-based data in Canada can be challenging.[8,40,41] Other racialized populations within high-income countries will be excluded, as they have their own distinct identities and face unique inequities; therefore, a separate ScR, to which they are the primary focus, may be more appropriate.[8]

The concept (C) of determinants of vaccine hesitancy encompasses reasons for unwillingness to be vaccinated and other similarly meaning sentiments. Due to the increasing trend of vaccine hesitancy, measures should be taken to identify and understand underlying factors and to inform the creation of effective and targeted solutions.[18,19,42] In this ScR, attempts will be made to explore all relevant concepts and their boundaries, with the only exclusion pertaining to the dates of publication. Publication dates will be restricted to December 2020 to current, to provide contemporary evidence sources that are also inclusive to the public availability of the COVID-19 vaccine.[3,18,19,43]

The context (C) of public health has no exclusion criteria; both vulnerable populations and vaccinations are a primary concern of public health. Despite the ambiguity of some terms used within the research question, these are consistent with Arkey and O’Malley,[44] which recommends maintaining a wide approach to generate a breadth of literature; more parameters could be added once a general scope and volume of evidence sources is obtained.

### Planned Approach to Evidence Searching, Selection, and Data Extraction

The intention of this section is to provide a transparent and auditable search strategy and to provide structure to the proposed ScR.[32] The search strategy can be found in S1 File.

### Planned Approach to Evidence Searching

A comprehensive search strategy will be developed to identify relevant evidence sources based on the research questions and PCC framework. To increase the breadth of the ScR, source evidence will be open to all types, including primary studies, secondary studies, grey literature, poster presentations, abstracts, reports, and so on. Only French and English evidence sources will be used due to feasibility reasons, as these are the primary languages of the researchers, and translation is not available. In addition, only evidence sources with a primary focus on African, Caribbean, and/or Black populations will be included so that issues specific to this target population are addressed. Furthermore, the time frame will be restricted to 2020-current (July 19^th^, 2022) to provide contemporary findings, given the dynamic nature of vaccine uptake, particularly during the COVID-19 pandemic. All evidence sources from the searches will be recorded in the Preferred Reporting Items for Systematic Reviews (PRISMA) flowchart adapted from Page et al.[45]

The search strategy will follow the three-step process recommended by JBI[32]; step 1 will involve an initial search to identify a list of terms; step 2 will be the implementation of the search strategy based on the identified terms, and step 3 will involve a hand search of the references from selected evidence sources, direct contacting of authors were performed due to time limitations.

### Step 1: Initial Search to Identify the List of Relevant Terms

To help capture the breadth of low vaccine uptake in ACB populations relative to public health in high-income countries, a piloting of keywords and Medical Subject Headings (MeSH) terms were conducted by research team members SB and JE with information specialist VC June 2022. Initial searches were used to inform the iterative process of the ScR, by potentially refining and allowing for new sources and keywords to be added.[32]

Search strategies will be developed by an information specialist (VC) and peer-reviewed using the PRESS guideline.[46] The search will be conducted in: MEDLINE(R) ALL (OvidSP), Embase (OvidSP), CINAHL (EBSCOHost), APA PsycInfo (OvidSP), Cochrane Central Register of Controlled Trials (OvidSP), Cochrane Database of Systematic Reviews (OvidSP), Allied & Complimentary Medicine Database (Ovid SP), and Web of Science. Each database will be searched from its inception until July 19, 2022, for the concept of “Vaccine uptake” and “African, Black and Caribbean” populations using a combination of subject headings and keywords.

### Step 2: Implementation of Search Strategy Based on Identified Terms

Drafting the search strategy will be informed by two Cochrane reviews [47,48] and a protocol in the Open Science Framework [38] for the concept of vaccine uptake. The concept of African, Caribbean and Blacks will be informed by consulting Hope et al.’s [49] systematic review. No search filters or language limits will be used, and publication restrictions will be applied to the search.

### Step 3: Hand Searches and Reference List

Research team members (MA, PB, GO, KS, RS, ECO, ID, SJ, HO, and SB). will manually search the retrieved publications’ reference lists to find all relevant studies that they would consider. All references that satisfy the eligibility requirements will be checked against our initial list of articles for duplication, and any that are found to be duplicates will be deleted. Additional hand searches will not be done due to time constraints. All other sources of evidence will be listed in the PRISMA flowchart.

### The Planned Approach to Selection

All evidence sources from the database searches will be uploaded into the software program Covidence, which will remove duplicate articles. They will then be screened by their title and abstract based on the aforementioned eligibility criteria by two members of the research team (MA, PB, GO, RS, ECO, ID, SJ, SB) being resolved by a third team member. During this process, any additional duplicate articles found will be removed. Articles will be selected based on the title, and abstract screening, followed by the full text by two research team members (MA, PB, GO, KS, RS, ECO, ID, SJ, SB), and a third member will resolve any conflicts. The number of excluded articles will be recorded in the PRISMA flowchart with the rationale for their exclusion.

### The Planned Approach to Data Extraction

A critical appraisal of the selected evidence sources will not be performed; it is not typically performed in an ScR.[32] This helps maintain a breadth of evidence from various sources; including, primary studies, secondary studies, grey literature, websites, blogs, reports etc.

However, risk for bias is created by not critically appraising the evidence sources, and thus findings will not be synthesized.[32] However, in order to create a descriptive narrative of the evidence sources, an extraction table will be created to provide an analytical framework.[44]

To calibrate the table, reviewers will independently extract three evidence sources, which will subsequently be compared and, any discrepancies between the reviewers will be discussed until at least a 75% agreement is achieved. At this point, any remaining issues will be recorded, and the full extraction of all sources of evidence will occur.[32] Creating the table is an iterative process, and modifications may be made as through the same process as more evidence emerges.[50] Refer to S2 File for a template of the extraction table; any further changes made will be documented and a rationale will be provided. The template for this table contains general information and information that can help inform the research questions and objectives; namely, first author, date, country of origin, population, vaccination type, study design, sample size, findings, and conclusion. Given that several different types of evidence sources are anticipated, it is not expected that all categories in the general extraction table will be completed for each evidence source; not applicable (N/A) will be written, where appropriate. However, the extracted data from each evidence source will align with the research question and objectives based on reviewing its entire text. Microsoft Word will be used during the extraction process to record and organize the data. The extracted information will be used to inform the collation and summarization of the findings from the evidence sources.

### Planned Approach to Collation and Summarization and Presentation of Evidence

Thematic mapping (TM) [51] will be used to collate and summarize the extracted data, as it is ideal for accommodating the multiple types of evidence sources in this study. This method also allows for the production of themes while maintaining the breadth of information within each theme through the creation of subthemes (descriptive and analytical themes), which is consistent with Arkey and O’Malley,[44] which has stated that a ScR seeks to explore the breadth of existing evidence sources and not to qualify the evidence or provide generalizable robust findings and Peters et al.[32] which has stated that results should be descriptively mapped rather than synthesized. The critical appraisal step in TM will not be performed, as it is unnecessary for a ScR.

### Thematic Mapping is a three-phase process

**Figure 1.**
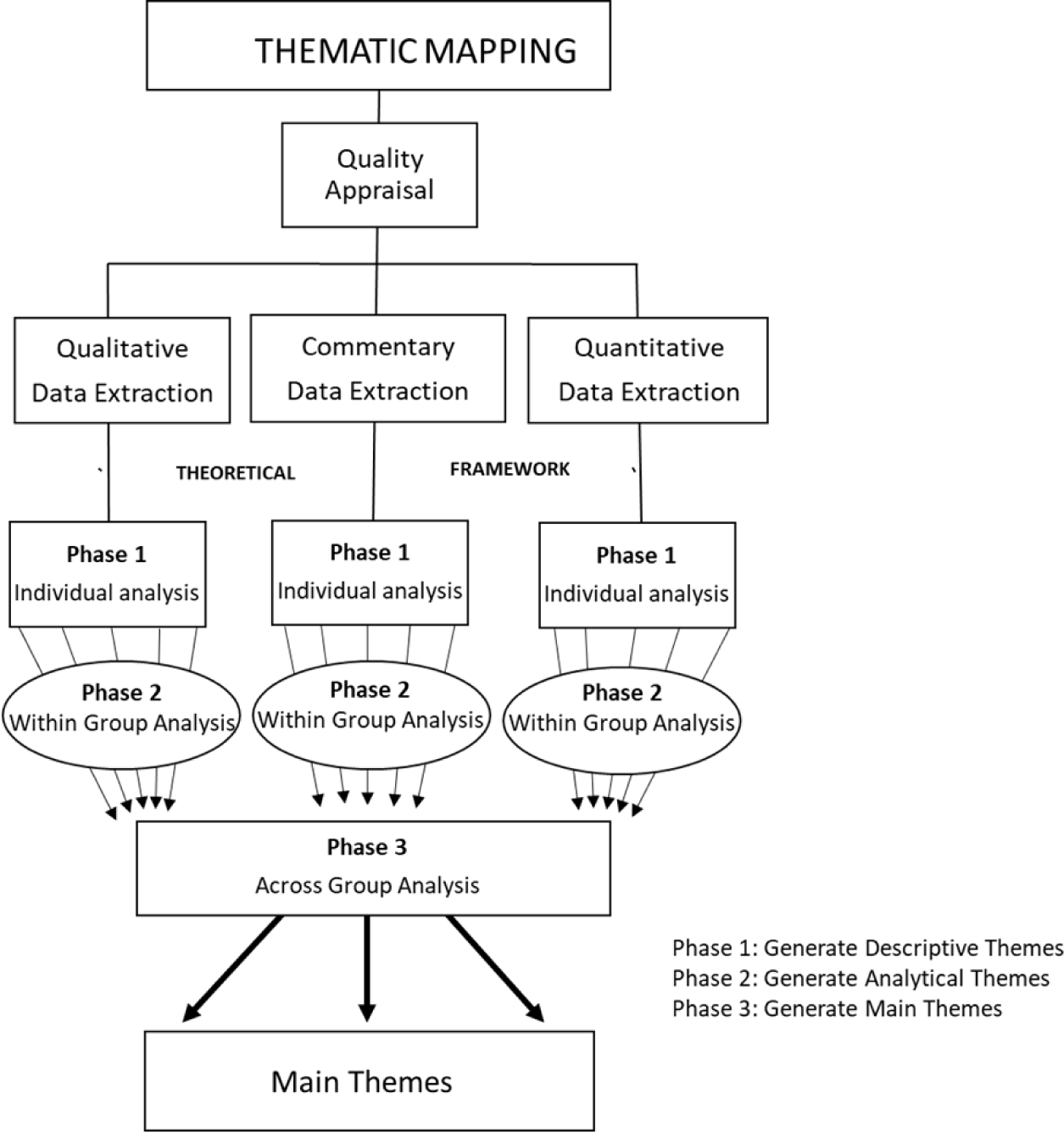
Thematic mapping. This diagram from Etowa et al.[51] illustrates the steps in thematic mapping. These steps will be followed in our study with the exception of the qualitative appraisal which is not required in a scoping review.[32]

The aim of Phase 1 will be to create initial codes and descriptive themes which are informed by the theoretical underpinnings used in this study; namely, intersectionality, the social determinants of health (SDOH), and the socio-economic model (SEM) as they relate to low vaccine uptake in African, Caribbean, and Black (ACB) populations relative to public health in high-income countries. In this phase, the articles will be divided by article types. To ensure methodological rigor two reviewers will independently code the extracted data from each article; namely, MA and SB will extract the qualitative articles, GO and SB will extract the quantitative articles, KS and SB will extract the commentaries, and RS and SB will extract the mixed methods articles, any conflicts will be resolved by a third researcher JB. Consistent with Objective 1 of this study, which is to identify concepts and boundaries of existing evidence sources on low vaccine uptake in ACB populations, the generated codes from each type of article will be used to create descriptive themes which will then further define the context and boundaries of concepts within each group. The descriptive themes will be based on grouping the codes by similarities and differences until a consensus was reached.

Within Phase 2, analytical themes will be created to gain further insight into the characteristics of the descriptive themes that reflected the content within each article grouping (qualitative, quantitative, mixed method and commentaries) separately. The descriptive themes from each article will be grouped based on similarities and differences into analytical themes through induction and interpretation that is consistent with the PCC research question and the theoretical underpinnings as in Phase 1. The iterative process of creating the final analytical themes will conclude when a consensus was reached between the research team members.

Within Phase 2, main themes will be created to give a broad overview of findings based on the similarities and differences across all groups (article types). These main themes will be used to map the evidence on the concepts and boundaries, identify gaps in the research (Objective 2), and determine existing interventions to improve low vaccine uptake in the study population (Objective 3).

A descriptive narrative summary will be used to describe these findings because an ScR’s analysis is descriptive by nature.[32]

### Ethics Approval and Consent to Participate

This study will use published evidence sources with no human or animal participants; thus ethical approval and consent to participate will not be applicable.

## Discussion

In order to limit bias, this ScR procedure will predefine the search question, objectives, methodology, eligibility criteria, search methods, data extraction techniques, summary, and the presentation of anticipated findings.[32] This ScR will provide a broad perspective from a variety of evidence sources, with a focus on understanding the circumstances and limitations of ACB communities’ low vaccination rates. Moreover, it will aid in determining the range of available evidence sources available and identify research gaps. This ScR will gain insight into different vulnerable populations by pinpointing the factors that result in low vaccine uptake in ACB populations and will offer insight into the policies, practices, and research that can be used to meet the needs of ACB and other vulnerable populations.

## Strengths and Limitations

The anticipated broad selection and types of evidence sources will include grey literature, to maximize the breadth of findings thus, strengthening this scoping review. However, the dynamic nature of vaccination hesitancy, which will necessitate surveillance and acquiring new information to remain current, limits this study. Another restriction that could result in the loss of necessary data is the language limitation to English and French. Another drawback is that the evidence sources might not be critically appraised, which might impact the study’s validity, ability to be validated, and ability to synthesize and analyze the extracted findings.

Despite the limitations, this scoping review will help to identify, define, and highlight inequities that may impact health behaviors associated with low vaccine uptake, because it strives to increase the understanding of factors that determine low vaccine uptake in ACB populations. It is anticipated that this ScR will improve service provider knowledge in this respect and will contribute to the creation of adequate interventions.

## Dissemination

The results from this scoping review will be disseminated through peer-reviewed open-access journals and conferences targeting public health stakeholders, vaccination campaigns and overcoming inequities in healthcare.

## Supporting Information

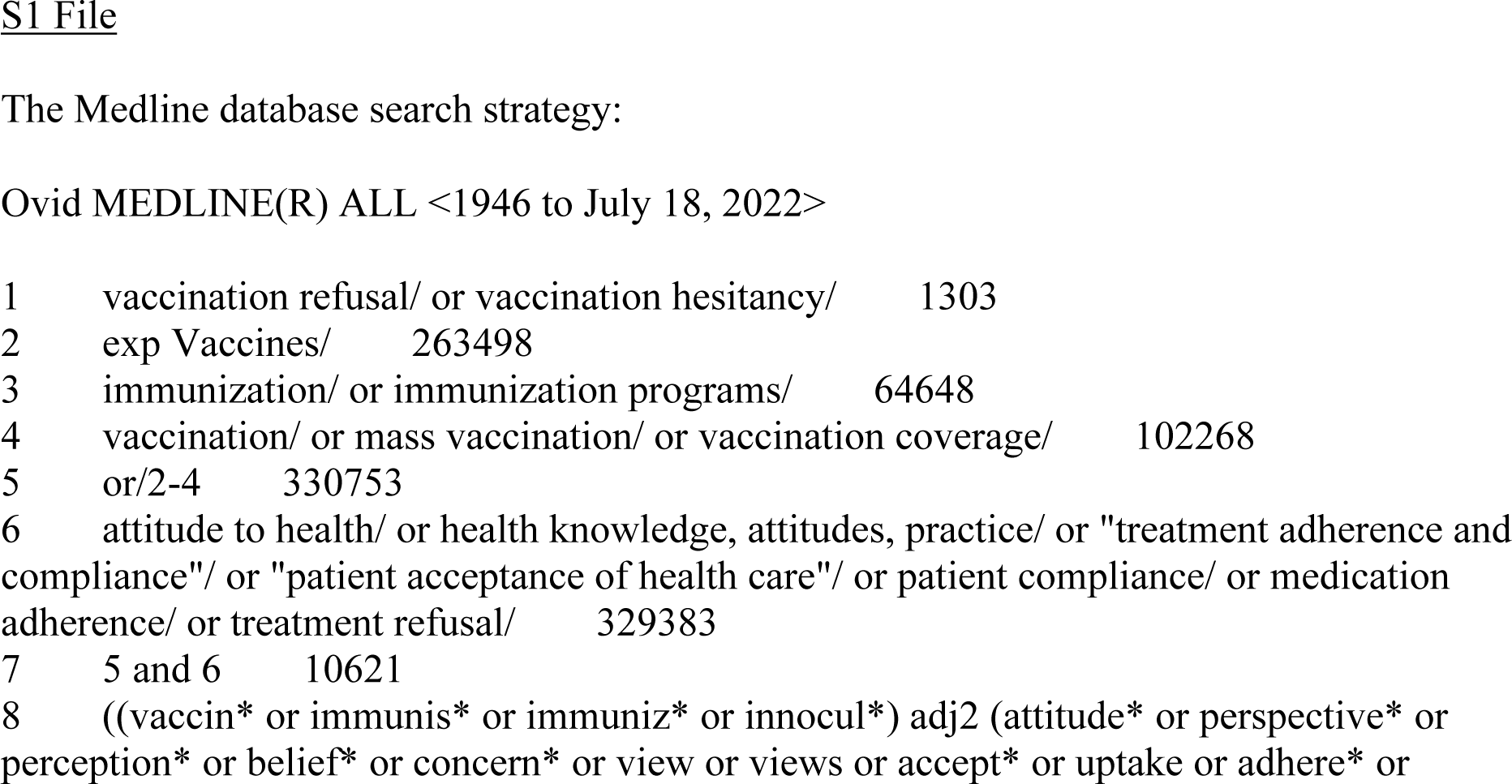
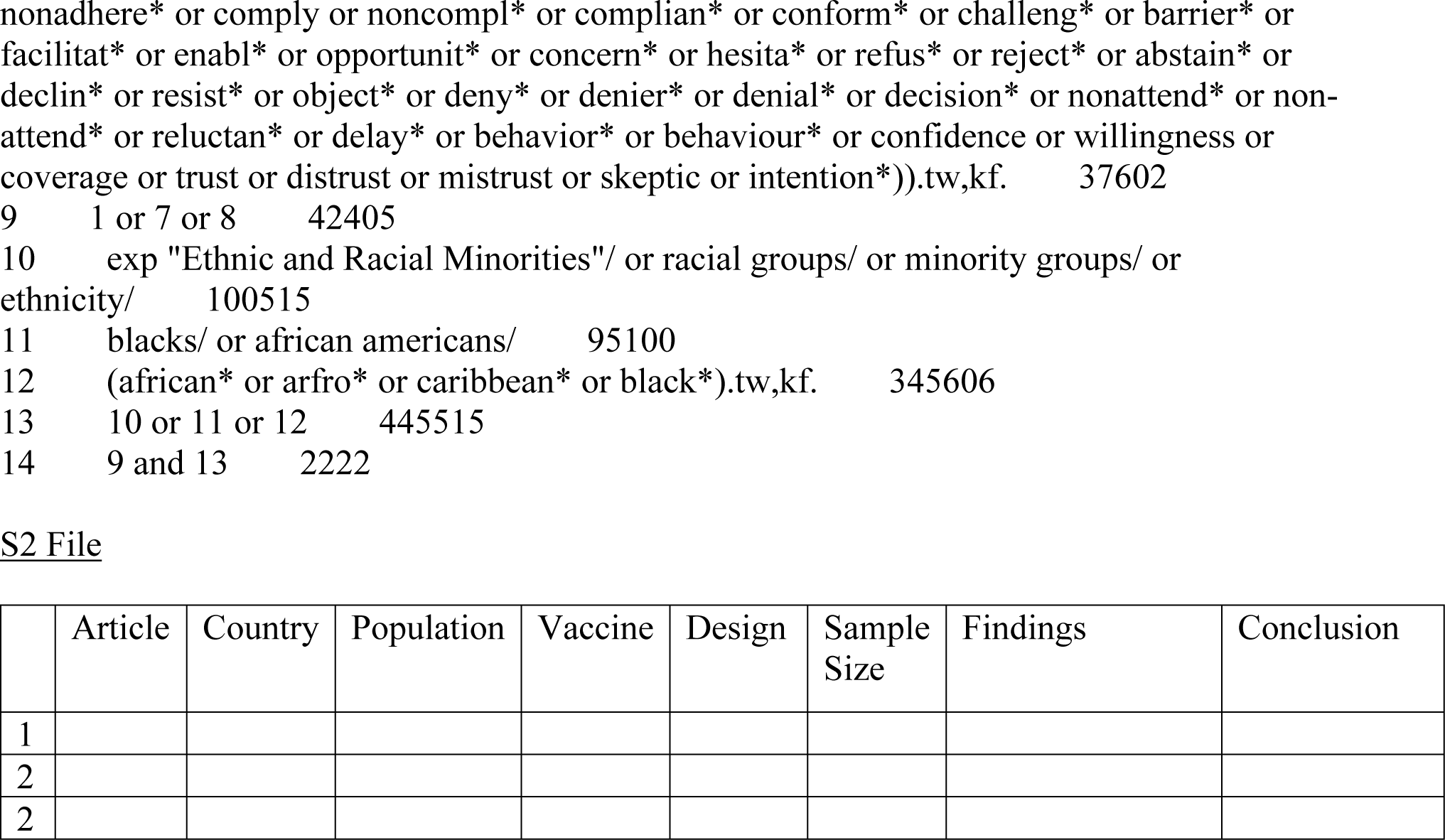

## Data Availability

No datasets were generated or analysed during the current study. All relevant data from this study will be made available upon study completion.

## Acknowledgements

We thank Nigèle Langlois, MLIS (Research Librarian, University of Ottawa Library) for peer review of the MEDLINE search strategy.

## References

1. The Canadian COVID-19 Genomics Network (CanCOGeN). Understanding the impact of COVID-19 on Black Canadians. In: GenomeCanada [Internet]. 26 Feb 2021 [cited 10 Jan 2023]. Available: https://genomecanada.ca/understanding-impact-covid-19-black-canadians/

2. Eissa A, Lofters A, Akor N, Prescod C, Nnorom O. Increasing SARS-CoV-2 vaccination rates among Black people in Canada. CMAJ. 2021;193: E1220–E1221. doi:10.1503/cmaj.210949

3. Statistics Canada. COVID-19 vaccine willingness among Canadian population groups. 26 Mar 2021 [cited 10 Jan 2023]. Available: https://www150.statcan.gc.ca/n1/pub/45-28-0001/2021001/article/00011-eng.htm

4. Kricorian K, Turner K. COVID-19 Vaccine Acceptance and Beliefs among Black and Hispanic Americans. PLOS ONE. 2021;16: e0256122. doi:10.1371/journal.pone.0256122

5. Public Health Agency of Canada. CPHO Sunday Edition: The Impact of COVID-19 on Racialized Communities. 21 Feb 2021 [cited 10 Jan 2023]. Available: https://www.canada.ca/en/public-health/news/2021/02/cpho-sunday-edition-the-impact-of-covid-19-on-racialized-communities.html

6. Dalsania AK, Fastiggi MJ, Kahlam A, Shah R, Patel K, Shiau S, et al. The Relationship Between Social Determinants of Health and Racial Disparities in COVID-19 Mortality. J Racial and Ethnic Health Disparities. 2022;9: 288–295. doi:10.1007/s40615-020-00952-y

7. Dryden O, Nnorom O. Time to dismantle systemic anti-Black racism in medicine in Canada. CMAJ. 2021;193: E55–E57. doi:10.1503/cmaj.201579

8. Thompson E, Edjoc R, Atchessi N, Striha M, Gabrani-Juma I, Dawson T. COVID-19: A case for the collection of race data in Canada and abroad. CCDR. 2021;47: 300–304. doi:10.14745/ccdr.v47i78a02

9. Shafiq M, Elharake JA, Malik AA, McFadden SM, Aguolu OG, Omer SB. COVID-19 Sources of Information, Knowledge, and Preventive Behaviors Among the US Adult Population. Journal of Public Health Management and Practice. 2021;27: 278. doi:10.1097/PHH.0000000000001348

10. Acharya A, Lam K, Danielli S, Ashrafian H, Darzi A. COVID-19 vaccinations among Black Asian and Minority Ethnic (BAME) groups: Learning the lessons from influenza. International Journal of Clinical Practice. 2021;75: e14641. doi:10.1111/ijcp.14641

11. Olanipekun T, Effoe VS, Olanipekun O, Igbinomwanhia E, Kola-Kehinde O, Fotzeu C, et al. Factors influencing the uptake of influenza vaccination in African American patients with heart failure: Findings from a large urban public hospital. Heart & Lung. 2020;49: 233–237. doi:10.1016/j.hrtlng.2019.12.003

12. Marsh HA, Malik F, Shapiro E, Omer SB, Frew PM. Message Framing Strategies to Increase Influenza Immunization Uptake Among Pregnant African American Women. Matern Child Health J. 2014;18: 1639–1647. doi:10.1007/s10995-013-1404-9

13. World Health Organization (WHO). Ten threats to global health in 2019. [cited 10 Jan 2023]. Available: https://www.who.int/news-room/spotlight/ten-threats-to-global-health-in-2019

14. Strategic Advisory Group of Experts (SAGE). Report of the Sage Working Group on Vaccine Hesitancy. In: The Compass for SBC [Internet]. 2014 [cited 12 Jan 2023]. Available: https://thecompassforsbc.org/sbcc-tools/report-sage-working-group-vaccine-hesitancy

15. Dubé E, Laberge C, Guay M, Bramadat P, Roy R, Bettinger JA. Vaccine hesitancy. Human Vaccines & Immunotherapeutics. 2013;9: 1763–1773. doi:10.4161/hv.24657

16. Koons C. When Nurses Refuse to Get Vaccinated. Bloomberg.com. 28 Aug 2021. Available: https://www.bloomberg.com/news/newsletters/2021-08-28/when-nurses-refuse-to-get-vaccinated. Accessed 10 Jan 2023.

17. Bhagavathula AS, Aldhaleei WA, Rahmani J, Mahabadi MA, Bandari DK. Knowledge and Perceptions of COVID-19 Among Health Care Workers: Cross-Sectional Study. JMIR Public Health Surveill. 2020;6: e19160. doi:10.2196/19160

18. Harrison EA, Wu JW. Vaccine confidence in the time of COVID-19. Eur J Epidemiol. 2020;35: 325–330. doi:10.1007/s10654-020-00634-3

19. World Health Organization (WHO). Immunization coverage. 2021a [cited 10 Jan 2023]. Available: https://www.who.int/news-room/fact-sheets/detail/immunization-coverage

20. Mayo Clinic. Herd immunity and COVID-19: What you need to know. In: Mayo Clinic [Internet]. 2021 [cited 10 Jan 2023]. Available: https://www.mayoclinic.org/diseases-conditions/coronavirus/in-depth/herd-immunity-and-coronavirus/art-20486808

21. Government of Canada (GC). Canadian Immunization Guide: Introduction. 2020 [cited 10 Jan 2023]. Available: https://www.canada.ca/en/public-health/services/canadian-immunization-guide/introduction.html

22. Oku A, Oyo-Ita A, Glenton C, Fretheim A, Ames H, Muloliwa A, et al. Communication strategies to promote the uptake of childhood vaccination in Nigeria: a systematic map. Global Health Action. 2016;9: 30337. doi:10.3402/gha.v9.30337

23. Center for Disease Control and Prevention (CDC). Overview, History, and How It Works | CDC. 9 Sep 2020 [cited 10 Jan 2023]. Available: https://www.cdc.gov/vaccinesafety/ensuringsafety/history/index.html

24. Nguyen TC, Gathecha E, Kauffman R, Wright S, Harris CM. Healthcare distrust among hospitalised black patients during the COVID-19 pandemic. Postgraduate Medical Journal. 2022;98: 539–543. doi:10.1136/postgradmedj-2021-140824

25. Shen SC, Dubey V. Addressing vaccine hesitancy: Clinical guidance for primary care physicians working with parents. Can Fam Physician. 2019;65: 175–181.

26. World Health Organization (WHO). COVID-19 Global Risk Communication and Community Engagement Strategy – interim guidance. 2021b [cited 10 Jan 2023]. Available: https://www.who.int/publications-detail-redirect/covid-19-global-risk-communication-and-community-engagement-strategy

27. Bogart LM, Ojikutu BO, Tyagi K, Klein DJ, Mutchler MG, Dong L, et al. COVID-19 Related Medical Mistrust, Health Impacts, and Potential Vaccine Hesitancy Among Black Americans Living With HIV. J Acquir Immune Defic Syndr. 2021;86: 200–207. doi:10.1097/QAI.0000000000002570

28. Waisel DB. Vulnerable populations in healthcare. Current Opinion in Anesthesiology. 2013;26: 186. doi:10.1097/ACO.0b013e32835e8c17

29. Office for National Statistics. Coronavirus and Vaccine Hesitancy, Great Britain: 31 March to 25 April 2021. 2021 [cited 22 Feb 2023]. Available: https://www.ons.gov.uk/peoplepopulationandcommunity/healthandsocialcare/healthandwellbeing/datasets/coronavirusandvaccinehesitancygreatbritain

30. Tricco AC, Lillie E, Zarin W, O’Brien K, Colquhoun H, Kastner M, et al. A scoping review on the conduct and reporting of scoping reviews. BMC Medical Research Methodology. 2016;16: 15. doi:10.1186/s12874-016-0116-4

31. Lockwood C, dos Santos KB, Pap R. Practical Guidance for Knowledge Synthesis: Scoping Review Methods. Asian Nursing Research. 2019;13: 287–294. doi:10.1016/j.anr.2019.11.002

32. Peters M DJ, Godfrey C, McInerney P, Munn Z, Tricco AC, Khalil H. Chapter 11: Scoping reviews - JBI Manual for Evidence Synthesis - JBI Global Wiki. 2020 [cited 10 Jan 2023]. Available: https://synthesismanual.jbi.global.

33. Munn Z, Peters MDJ, Stern C, Tufanaru C, McArthur A, Aromataris E. Systematic review or scoping review? Guidance for authors when choosing between a systematic or scoping review approach. BMC Medical Research Methodology. 2018;18: 143. doi:10.1186/s12874-018-0611-x

34. Kamal A, Hodson A, Pearce JM. A Rapid Systematic Review of Factors Influencing COVID-19 Vaccination Uptake in Minority Ethnic Groups in the UK. Vaccines. 2021;9: 1121. doi:10.3390/vaccines9101121

35. Biswas MR, Alzubaidi MS, Shah U, Abd-Alrazaq AA, Shah Z. A Scoping Review to Find Out Worldwide COVID-19 Vaccine Hesitancy and Its Underlying Determinants. Vaccines. 2021;9: 1243. doi:10.3390/vaccines9111243

36. Yasmin F, Najeeb H, Moeed A, Naeem U, Asghar MS, Chughtai NU, et al. COVID-19 Vaccine Hesitancy in the United States: A Systematic Review. Frontiers in Public Health. 2021;9. Available: https://www.frontiersin.org/articles/10.3389/fpubh.2021.770985

37. Salomoni MG, Di Valerio Z, Gabrielli E, Montalti M, Tedesco D, Guaraldi F, et al. Hesitant or Not Hesitant? A Systematic Review on Global COVID-19 Vaccine Acceptance in Different Populations. Vaccines. 2021;9: 873. doi:10.3390/vaccines9080873

38. Thota AB. PROTOCOL: Scoping Review of Interventions to Increase Vaccination Uptake for Racial and Ethnic Minorities and Indigenous Population Groups Living in High-Income Countries. OSF Preprints; 2022. doi:10.31219/osf.io/t3ykz

39. World Bank. High income. 2022 [cited 9 Jun 2022]. Available: https://data.worldbank.org/country/XD

40. Annett C. Inequities in COVID-19 Health Outcomes: The Need for Race- and Ethnicity- Based Data. In: HillNotes [Internet]. 8 Dec 2020 [cited 11 Jan 2023]. Available: https://hillnotes.ca/2020/12/08/inequities-in-covid-19-health-outcomes-the-need-for-race-and-ethnicity-based-data/

41. Government of Ontario. Ontario’s Regulatory Registry. Government of Ontario, Ministry of Economic Development, Job Creation and Trade; 2021 [cited 11 Jan 2023]. Available: https://www.ontariocanada.com/registry/view.do?postingId=32967&language=en

42. Kumar D, Chandra R, Mathur M, Samdariya S, Kapoor N. Vaccine hesitancy: understanding better to address better. Israel Journal of Health Policy Research. 2016;5: 2. doi:10.1186/s13584-016-0062-y

43. The Guardian. Covid: UK woman who was first in world to receive vaccine has second dose. The Guardian. 29 Dec 2020. Available: https://www.theguardian.com/world/2020/dec/29/first-person-to-get-covid-jab-receives-follow-up-dose. Accessed 11 Jan 2023.

44. Arksey H, O’Malley L. Scoping studies: towards a methodological framework. International Journal of Social Research Methodology. 2005;8: 19–32. doi:10.1080/1364557032000119616

45. Page MJ, McKenzie JE, Bossuyt PM, Boutron I, Hoffmann TC, Mulrow CD, et al. The PRISMA 2020 statement: an updated guideline for reporting systematic reviews. BMJ. 2021;372: n71. doi:10.1136/bmj.n71

46. McGowan J, Sampson M, Salzwedel DM, Cogo E, Foerster V, Lefebvre C. PRESS Peer Review of Electronic Search Strategies: 2015 Guideline Statement. Journal of Clinical Epidemiology. 2016;75: 40–46. doi:10.1016/j.jclinepi.2016.01.021

47. Abdullahi LH, Kagina BM, Ndze VN, Hussey GD, Wiysonge CS. Improving vaccination uptake among adolescents. Cochrane Database of Systematic Reviews. 2020 [cited 18 Jan 2023]. doi:10.1002/14651858.CD011895.pub2

48. Cooper S, Schmidt B-M, Sambala EZ, Swartz A, Colvin CJ, Leon N, et al. Factors that influence parents’ and informal caregivers’ views and practices regarding routine childhood vaccination: a qualitative evidence synthesis. Cochrane Database of Systematic Reviews. 2021 [cited 18 Jan 2023]. doi:10.1002/14651858.CD013265.pub2

49. Hope MO, Taggart T, Galbraith-Gyan KV, Nyhan K. Black Caribbean Emerging Adults: A Systematic Review of Religion and Health. J Relig Health. 2020;59: 431–451. doi:10.1007/s10943-019-00932-5

50. Tricco AC, Lillie E, Zarin W, O’Brien KK, Colquhoun H, Levac D, et al. PRISMA Extension for Scoping Reviews (PRISMA-ScR): Checklist and Explanation. Ann Intern Med. 2018;169: 467–473. doi:10.7326/M18-0850

51. Etowa J, Demeke J, Abrha G, Worku F, Ajiboye W, Beauchamp S, et al. Social determinants of the disproportionately higher rates of COVID-19 infection among African Caribbean and Black (ACB) population: A systematic review protocol. J Public Health Res. 2021;11: 2274. doi:10.4081/jphr.2021.2274

